# Validating quantitative PCR assays for cell-free DNA detection without DNA extraction: Exercise induced kinetics in systemic lupus erythematosus patients

**DOI:** 10.1101/2021.01.17.21249972

**Authors:** Elmo W.I. Neuberger, Alexandra Brahmer, Tobias Ehlert, Katrin Kluge, Keito F.A. Philippi, Simone C. Boedecker, Julia Weinmann-Menke, Perikles Simon

## Abstract

Circulating cell-free DNA (cfDNA) has been investigated as a screening tool for many diseases. To avoid expensive and time-consuming DNA isolation, direct quantification PCR assays can be established. However, rigorous validation is required to provide reliable data in the clinical and non-clinical context. Considering International Organization for Standardization, as well as bioanalytical method validation guidelines we provide a comprehensive procedure to validate assays for cfDNA quantification from unpurified blood plasma. A 90 and 222 bp assay was validated to study the kinetics of cfDNA after exercise in patients with systemic lupus erythematosus. The assays showed ultra-low limit of quantification (LOQ) with 0.47 and 0.69 ng/ml, repeatability ≤ 11.6% (95% CI: 8.1–20.3), and intermediate precision ≤ 12.1% (95% CI: 9.2-17.7). Incurred sample reanalysis confirmed the precision of the procedure. The additional consideration of pre-analytical factors shows that centrifugation speed and temperature do not change cfDNA concentrations. In SLE patients cfDNA increases ∼2 fold after all out walking exercise, normalizing after 60 min of rest. The established assays allow reliable and cost-efficient quantification of cfDNA in minute amounts of plasma in the clinical setting and can be used as a standard to control pre-analytical factors including cfDNA losses during purification.

## Introduction

Circulating cell-free DNA (cfDNA) is recognized to have reasonable prognostic or diagnostic potential in several pathological diseases including cancer, sepsis, and autoimmune diseases such as SLE^1,2^. In 1966 Tan et al. described high levels of cfDNA in serum of SLE patients^3^. Since that time, a plethora of studies confirmed elevated cfDNA concentrations in serum and plasma of SLE patients with active or inactive disease (reviewed in^2^). In patients with autoimmune disease, high levels of cfDNA are considered to be a relevant antigen for auto anti-body development^4–6^. Intriguingly, cfDNA concentrations increase about 10 fold during all out exercise, showing a half-life of ∼15 min in the healthy population^7–9^. While regular physical exercise is recommended for SLE patients, and several studies provided reasonable evidence that regular exercise reduces fatigue and increases cardiovascular fitness in SLE patients^10^, the kinetics of cfDNA during exercise in SLE patients are still unknown. A minimal invasive and cost-efficient assay is a valuable tool to monitor cfDNA concentrations, which might be indicative for overtraining or disease remission^11,12^.

Most cfDNA quantification assays require pre-analytical DNA extraction^13^. A process, which is costly, time consuming and has been shown to lead to variable loss of cfDNA depending on the extraction method employed^14,15^. To avoid cfDNA isolation, direct quantification assays have been established^14,16^. Using multilocus primers, which bind various sites in the human genome, a sufficient sensitivity can be reached. The hominoid specific long interspersed element 1 (LINE1) family 2 (L1PA2) is a suitable target, according to its abundance and specificity^17^.

A major prerequisite for reliable cfDNA detection is the evaluation of the assay performance. In 2019 the International Organization for Standardisation (ISO) published a guideline which comprehensively specifies the requirements for evaluating the performance of quantification methods for nucleic acid target sequences for quantitative real-time PCR (qPCR) and digital PCR (dPCR)^18^.

Based on previous work^19^, we describe the validation of reliable qPCRs assays for the quantification of cfDNA. Considering the relevant guidelines we emphasize the specificity, precision, including repeatability and intermediate precision, limit of detection (LOD), limit of quantification (LOQ), linearity, as well as incurred sample reanalysis. Since cfDNA typically shows a length of about 166bp^20^, the combination of a 90bp assay and 222bp assay enables the evaluation of DNA integrity^21,22^.

The assays could be applied successfully to quantify cfDNA samples in the clinical context. We show that exercise induced cfDNA increases start to decline after 30 min in SLE patients, normalizing after 60 - 90 min. Since elevated cfDNA concentrations have been discussed to possibly trigger enhanced inflammation or the production of anti-ds-DNA antibodies, low increases during and rapid decreases after exercise are rather positive aspects of cfDNA kinetics during exercise in SLE patients. The direct quantification enables a time and cost saving quantification assay, which further could be standard assay to control for pre-analytical factors including cfDNA losses during purification. Furthermore, according to its high sensitivity the assay can be used to study cfDNA in other body liquids, cell culture supernatant.

## Methods

### Ethics approval and exercise testing

SLE patients were recruited at the Department of internal Medicine, University Medical Center, Mainz, Germany. Patients with a stable immunosuppressive medical therapy and no active lupus nephritis were asked to participate in an exercise intervention study (ClinicalTrials.gov Identifier: NCT03942718, date of first registration 08.05.2019)^23^. The registered study aims to evaluate the effects of a 12-week exercise program on cardiorespiratory fitness. In the context of this assay validation study the samples from the initial examination time point were used to study the kinetics of cfDNA in SLE patients during and after acute exercise. The experimental procedures were approved by the Human Ethics Committee of Rhineland-Palatine and were in line with the Declaration of Helsinki of the World Medical Association. All participants gave their informed consent for study participation. A total of 28 eligible study participants performed a stepwise exercise test until volitional exhaustion with a modified walking protocol^23^. Speed and elevation of the treadmill were increased stepwise every three minutes until the participants stopped the test volitionally. Heart rate and respiratory gas exchange were monitored continuously.

### Blood sample collection

Venous blood samples from the cubital vein were collected before, directly after, and 90 min after the exercise test. The utilized blood collection products are listed in Supplementary Table S1. The samples were centrifuged immediately after drawing two times at 2500 x g for 15 min at room temperature (RT), to collect platelet-free plasma.^24^ Capillary blood samples were collected from the fingertip and stored at 4°C before centrifugation at 600 x g for 10 min. For comparison of centrifugation protocols, venous blood samples were centrifuged as indicated.

### Assay validation material

Linearity and accuracy of the direct quantification method was tested on a custom-made 401-bp fragment from the L1PA2 family (Supplementary Table S2). The fragment was synthesized by Eurofins MWG Operon, cloned into a pEX-A plasmid (Eurofins MWG Operon), amplified in bacteria before DNA isolation with QIAprep Spin Miniprep Kit (Qiagen, Hilden, Germany). After EcoRI restriction the 401 bp Fragment was gel purified with QIAquick Gel Extraction Kit (Qiagen, Hilden, Germany). The concentration was determined with a NanoDrop 3300 fluorospectrometer (Thermo Fisher Scientific, Inc., Waltham, MA) using Quant-iT™ PicoGreen dye (Thermo Fisher Scientific, Inc., Waltham, MA). For precision studies, a pool of venous plasma from six healthy subjects before (PRE exercise sample) and after exercise (POST exercise sample) was used.

### Sample preparation and qPCR

Plasma samples were diluted 1:10 in UltraPure™ DNase/RNase-Free H_2_O (Invitrogen, Waltham, MA). For the qPCR, 2 µl of the diluted plasma, were mixed with 12 µl master-mix, 1 µl primer mix, and measured in triplicates with a final volume of 5 µl. The primer sequences are listed in Supplementary Table S3. The final concentrations in the PCR reaction are 1.2X HiFi buffer (BioCat, Heidelberg, Germany), 0.3 mM of each dNTP (Carl Roth, Karlsruhe, Germany), 0.15X SYBR Green (Sigma, Taufkirchen, Germany), 0.04 IU Velocity Polymerase (BioCat, Heidelberg, Germany), and 140 nM of each primer. For amplification a CFX384 system with the following protocol was used: 98°C for 2 min followed by 35 cycles of 95°C for 10 sec (denaturation) and 64°C for 10 sec (annealing/extension) including the plate read. A melt curve from 70 – 95°C with 0.5 °C increments for 10 sec finished each run. If the triplicates of the samples show a SD Cq > 0.4, native plasma samples were re-diluted and reanalyzed.

### In silico calculation of L1PA2 copy numbers per genome

To determine the number of targets for the L1PA2_90bp and L1PA2_222bp primer pairs in the human genome (GRCh38/hg38), the UCSC Genome Browser *In-Silico PCR* tool was used (https://genome.ucsc.edu/cgi-bin/hgPcr). The Max Product Size (Maximum size of amplified region) was set to 660 bp, which equals the predicted maximal amplification rate of the polymerase during 10s amplification. Min Perfect Match (Number of bases that match exactly on 3’ end of primers) was set to 19 bp. The predicted sequences per chromosome and the length distribution are provided in Supplementary Fig. S4 and Table S5.

### Calculation of cfDNA concentration in plasma samples

To determine the cfDNA concentration in (ng/ml) from the Cq values of samples the following calculation is applied: ng/ml ≙pg/µl = 10^(Cq – intercept)/slope^ / 5µl / (1/75) * 3.23 pg / (3416 for L1PA2_90bp or 3237 for L1PA2_222bp). Using the slope and the intercept from the validated standard curve the total number of copies per 5µl qPCR reaction are calculated. A division by 5 results in copy numbers per µl. The copy numbers per µl plasma are calculated by considering the total dilution factor of plasma 1/75 (plasma dilution 1/10 and qPCR dilution: 2µl template per 15µl reaction). The copy number per µl is dived by the number of predicted sequences in the human genome (3416 for the L1PA2_90bp and 3237 for the L1PA2_222bp). The result reflects the number of genome equivalents (GE) per ml plasma. The resulting GE are multiplied with 3.23 pg (the expected weight of a haploid genome^25^) to calculate the pg concentration per µl plasma, equaling ng/ml.

### PCR run interplate calibration

The CFX384 real-time system calculates the C_q_ values based on the crossing point of the fluorescence traces with an auto-calculated threshold line. The threshold is dependent on the total number of samples and fluorescence intensity. To correct for inter-plate differences, two reference samples are measured in each run. For the analysis of the results, the threshold is adapted so that the mean of the C_q_ values of the reference samples equals the pre-defined mean C_q_ value. The pre-defined mean C_q_ value is based on the mean autocalculated C_q_ values of 10 PCR runs from two different operators. The indicated reference samples are a pre- and a post-exercise plasma samples from a single subject. Typical amplification curves are shown in Supplementary Fig. S6.

### Determination of assay performance

For the determination of the *dynamic range, assay linearity, LOD and LOQ* the purified L1PA2 DNA fragment was spiked into mouse plasma (1:10 final dilution). Mouse plasma is a relevant sample matrix and does not contain the human specific L1PA2 sequences. Three independent standard curves were prepared for each of the assays on different days. Each concentration (1 x 10^6^ - 25 copies per well) was pipetted in septet replicates (Fig. 1). H_2_O and mouse plasma NTC controls, as well as the reference samples were pipetted on each plate. For the determination of the LOQ the results from all standard curves were combined to generate imprecision profiles using the Variance Function Program VFP (v1.2) with R (v3.6.3) (Fig. 1). The LOD was defined as the lowest concentration of the target analyte which can be detected with a 95% detection rate^26,27^. Where applicable, the CLSI guideline EP17-A2 were used, defining a limit of blank (LOB) = Mean_copy number blank_ + 1.645 x SD_copy number blank_ and LOD = LOB + 1.645 x SD_low copy number sample_^28^.

**Figure 1.**
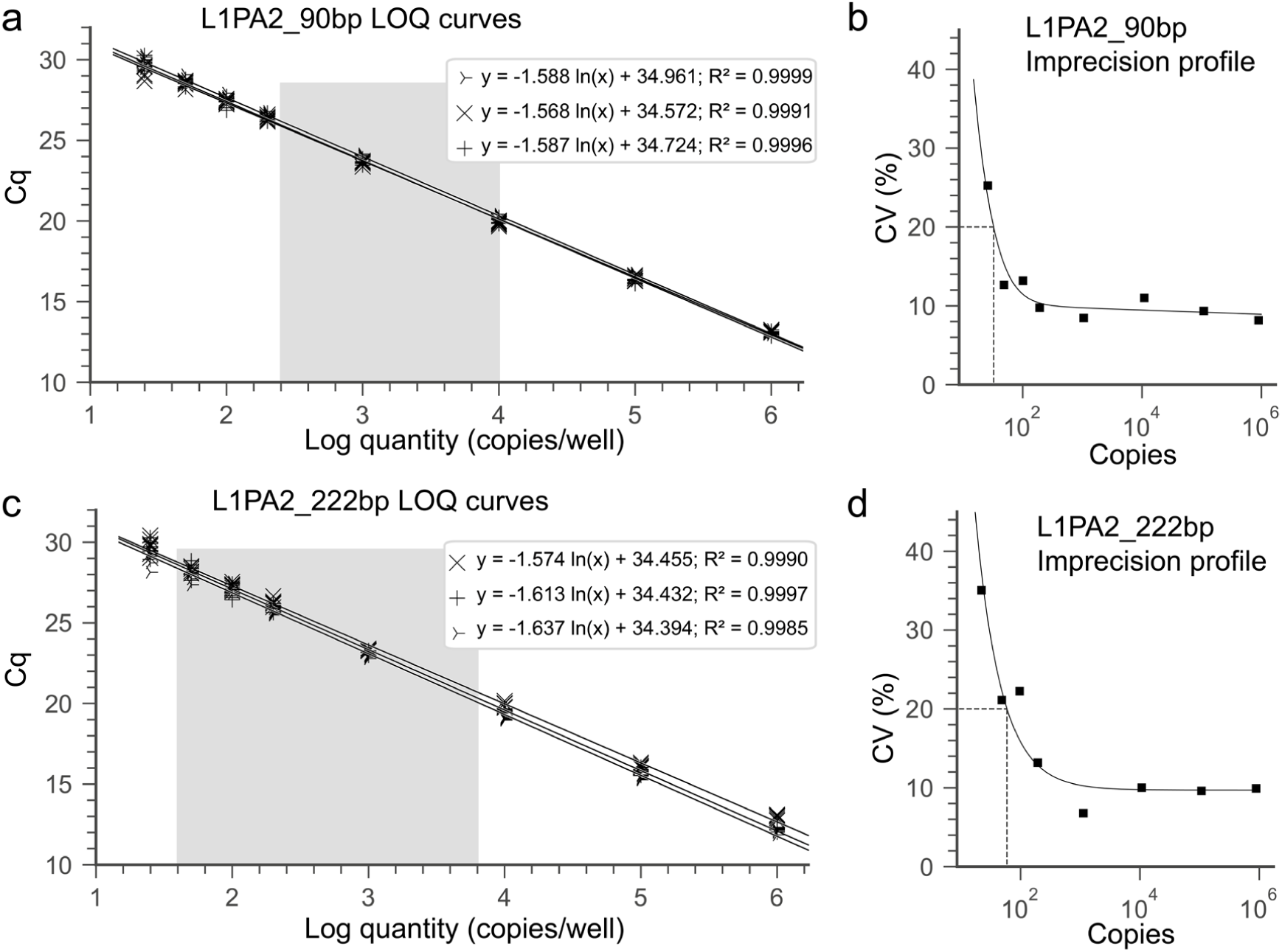
Standard curves and imprecision profiles for the L1PA2_90bp assay (**a, b**) and the L1PA2_222bp assay (**c, d**). Three independent standard curves, with septets per concentration were prepared for each assay. The imprecision profiles were calculated with the formulas: σ^2^ = 31.37 + 0.011 × U^1.975^ and σ^2^ = (6.10 + 0.097 × U)^2^ for the 90 bp (**b**) and 222 bp assay (**d**), respectively. The grey area indicates the quantification range of cfDNA concentrations in this study. Cq = cycle quantification value, LOQ = lower limit of quantification, CV = coefficient of variation.

#### Assay imprecision

To evaluate the influence of the plasma dilution process, 20 replicates of the PRE and POST samples were measured in a single run. 10 replicates were measured from the same diluted plasma pool and 10 replicates were diluted 1:10 uniquely. The coefficient of variation (CV) was calculated with the formula CV = (Standard Deviation / Mean) * 100. To estimate the repeatability and intermediate precision 20 replicates were run in duplicates over 10 different PCR runs.

#### Direct quantification compared to kit isolation

To compare the concentration of cfDNA before and after DNA isolation using custom DNA extraction kits, plasma was aliquoted in 15 tubes and frozen at -20°C. Five of the aliquots (1ml plasma) were purified with the QIAamp® Circulating Nucleic Acid kit (Qiagen, Hilden, Germany) (Purification kit 1), and the NucleoSnap cfDNA kit for cell-free DNA from plasma (Macherey-Nagel, Düren, Germany) (Purification kit 2), respectively. Additionally, 5 aliquots of 400µl plasma were isolated with the QIAamp DNA Blood Mini Kit (Qiagen, Hilden, Germany) (Purification kit3). All samples were eluted in H_2_O (1/10 of input volume). For the direct quantification, from each plasma aliquot a dilution was prepared. All samples were measured after a single freeze thaw cycle.

#### Incurred sample reanalysis

To verify the reliability of the cfDNA concentration in the study samples, a subset of two samples from each qPCR run was re-analyzed in the subsequent run. The percentage difference of the cfDNA concentration between the first and the repeated measurement is determined with the following equation: (Repeat – First) / Mean * 100, according to the Bioanalytical Method Validation Guidance for Industry^29^.

#### Storage stability

To determine the stability of plasma dilutions 20 samples of a former study were reanalyzed after > 2 month of storage at -20°C. To determine the storage stability of whole blood before centrifugation, four venous and capillary samples were taken consecutively and centrifuged at 2500 x g for 10 min directly after blood drawing, after 60 min, 90 min, or 180 min.

### Data analysis

The qPCR analysis was performed with the CFX Manager software Version: 3.1.1517.0823. Python (v3.7.6) with ‘matplotlib’ (v3.1.1), and ‘seaborn’ (v0.9.0), and R (v3.6.3) with ggplot2 (v3.3.2) were used for descriptive analysis. The imprecision calculation were performed using the R ‘VFP’ package (v1.2). Linear mixed models were conducted with the ‘lme4’ package (v1.1.23), to determine if capillary and venous cfDNA levels differ and respond differently to exercise. The ‘emmeans’ package (v1.4.8) was used to compute tukey corrected P values. Pearsons r or Spearman’s rho were calculated for normally or not normally distributed data, respectively.

## Results

### Characteristics of SLE patients

A total of 28 SLE patients (27 female, 1 male) were eligible for the study and participated in the performance diagnostic test without adverse events. The patients’ characteristics (Table 1) indicate a higher body mass index (BMI), and lower physical fitness (VO2peak) compared to the healthy population^30^.

**Table 1.** SLE patients’ characteristics. SLEDAI = Systemic Lupus Erythematosus Disease Activity Index, BMI = Body Mass Index.

| Characteristics of the study subjects | Mean | ± SD |
| --- | --- | --- |
| Age (years) | 49,42 | 8,70 |
| Weight (kg) | 77,53 | 18,54 |
| Height (cm) | 166,43 | 8,34 |
| BMI | 27,96 | 6,07 |
| SLEDAI | 6,50 | 4,42 |
| VO2peak (ml/min/kg) | 24,34 | 5,04 |
| Exercise time until exhaustion (min) | 13,61 | 3,67 |
| Medication, n (%) | 22 (78%) |  |
| Steroids | 17 (61%) |  |
| Hydrochloroquine | 14 (50%) |  |
| Belimumab | 4 (14%) |  |
| Cyclophosphamide | 1 (3%) |  |
| Azathioprine | 2 (7%) |  |
| Mycophenolate mofetil | 7 (25%) |  |
| No medication, n (%) | 6 (21%) |  |

### Assay performance

#### Dilution linearity and dynamic range

The independent LOQ curves show similar slope and y-intercept (Fig. 1). The efficiencies for the different standard curves are slightly lower than 90%. Ranging between 87.77 and 89.25% for the L1PA2_90bp assay and between 84.19 and 88.75% for L1PA2_222bp assay.

#### LOD and LOQ

All replicates of the low copy samples (25 copies) were detectable with the L1PA2_90bp and the L1PA2_222bp assay, not reaching LOD (Fig. 1). Of note, as shown in Supplemental Fig. S6, the NTCs of the L1PA2_90bp assay produce some background signal, which needs to be distinguishable from low copy samples. Since the data were normally distributed with homogenous variances, the LOB and LOQ were calculated according to the CLSI guideline^28^. The L1PA2_90bp assay shows a LOB of 8.39 copies and LOD of 18.59 copies. The LOQ, which was derived from the imprecision profile was 32.5 copies for the L1PA2_90bp and 59.26 copies for the L1PA2_222bp (Fig. 1), equaling a concentration of 0.47 ng/ml and 0.69 ng/ml, respectively.

#### Imprecision studies

The CVs were similar between the pooled or uniquely diluted plasma samples with 7.32% and 8.77% (PRE exercise sample), and 6.67% or 8.51% for the POST exercise sample (L1PA2_90bp assay) (Fig. 2a). For the L1PA2_222bp the CVs were 9.17 and 9.37% for the PRE exercise and 7.55 and 8.56 for the post exercise samples. The precision calculation for the measurements of the PRE and POST samples in repeated runs, resulted in a repeatability ≤ 11.59% (95% CI: 8.10 – 20.34) for the two assays.

**Figure 2.**
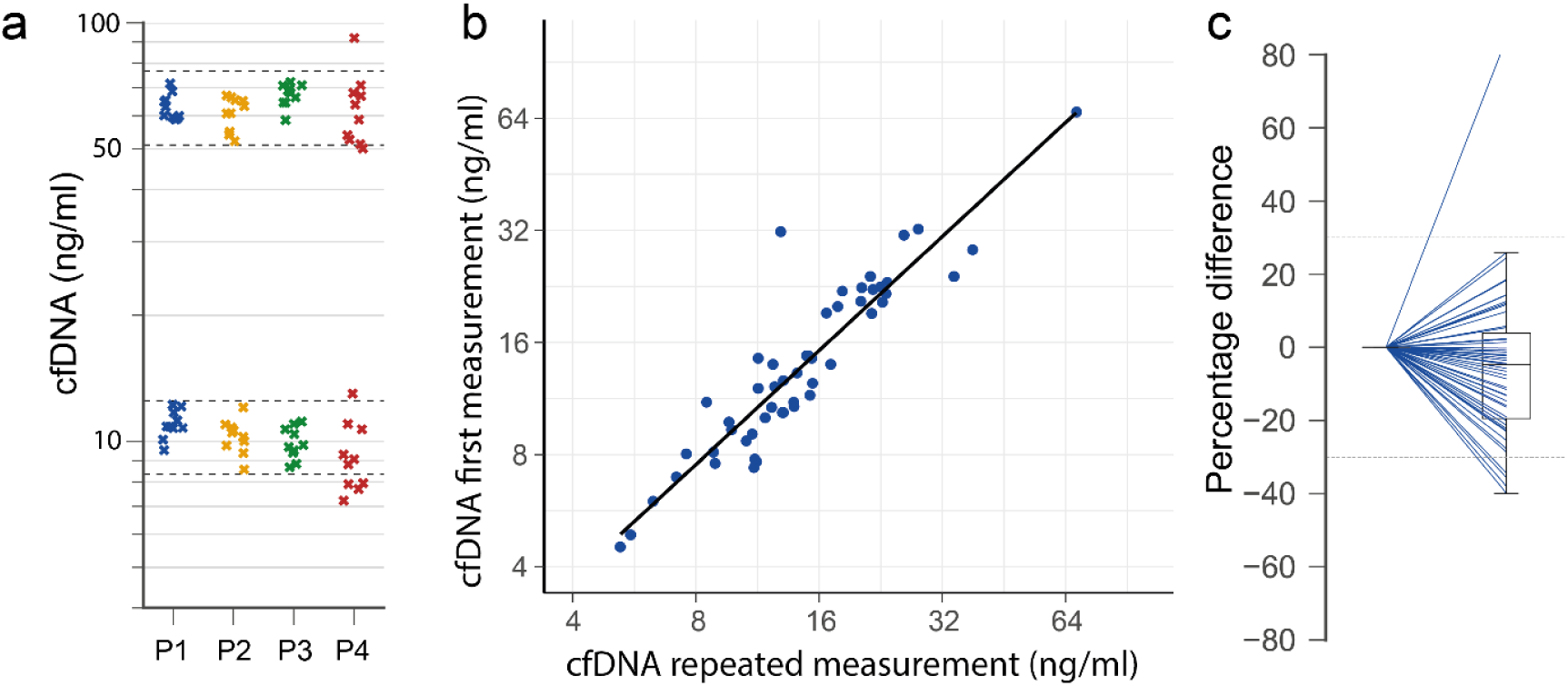
Precision of the L1PA2_90bp assay. (**a**) The same pool of plasma dilution of a low and high concentration sample was measured 10 times (P1), the same plasma was individually diluted 10 times (P2), the plasma dilution was measured in 10 consecutive runs in duplicate (P3 = mean of duplicates after inter-plate calibration, P4 = mean of duplicates before inter-plate calibration). The dashed lines indicate ± 20% of the nominal mean of conditions P1-3. Incurred sample reanalysis. From each run of the study samples, two samples were re-analyzed. (**b**) Correlation of the first and second measurement. (**c**) Percentage difference between original and second measurement.

#### Inter-run calibration

After the inter-run calibration the intermediate precision, calculated from 10 consecutive qPCR runs, reduced for the L1PA2_90bp assay from 23.80% (95% CI: 17.96 – 35.31) to 11.62% (95% CI: 8.77 – 17.22) (PRE sample), and from 24.75% (96% CI: 18.47 – 37.51) to 8.46% (95% CI: 6.40 – 12.45) (POST sample) (Fig. 2a). For the L1PA2_222bp assay the intermediate precision reduced for the PRE sample from 18.01% (95% CI: 12.58 – 31.61) to 12.97 (95% CI: 8.90-18.24), and the POST sample from 21.54% (95% CI: 16.04 – 32.78) to 12.06% (95% CI: 9.16 – 17.69).

### Stability of plasma dilutions

Plasma samples can be stored at -20°C or -80°C for years without affecting cfDNA concentration^31^. To determine the stability of 1:10 plasma dilutions, 20 diluted samples were re-analyzed with the L1PA2_90bp assay after storage for > 2 month at -20°C. cfDNA concentrations did not differ significantly (P = 0.35) and no degradation was obvious after a repeated freeze-thaw cycle. Prolonged storage of whole blood (up to 180 min) before centrifugation did not lead to relevant cfDNA concentration changes (Supplementary Fig. S7).

### Effects of cfDNA purification

The use of specific DNA isolation kits for cfDNA isolation from plasma (Purification kit 1 and 2) slightly affects the cfDNA concentration and DNA integrity (Fig. 3a). During the isolation process about 12% (± 9.5) and 23% (± 6.2) are lost with Purification kit 1 or 2, respectively. A kit for whole blood DNA isolation (Purification kit 3) leads to relevant DNA losses of about 84% and increased DNA integrity, which indicates a loss of short DNA fragments.

**Figure 3.**
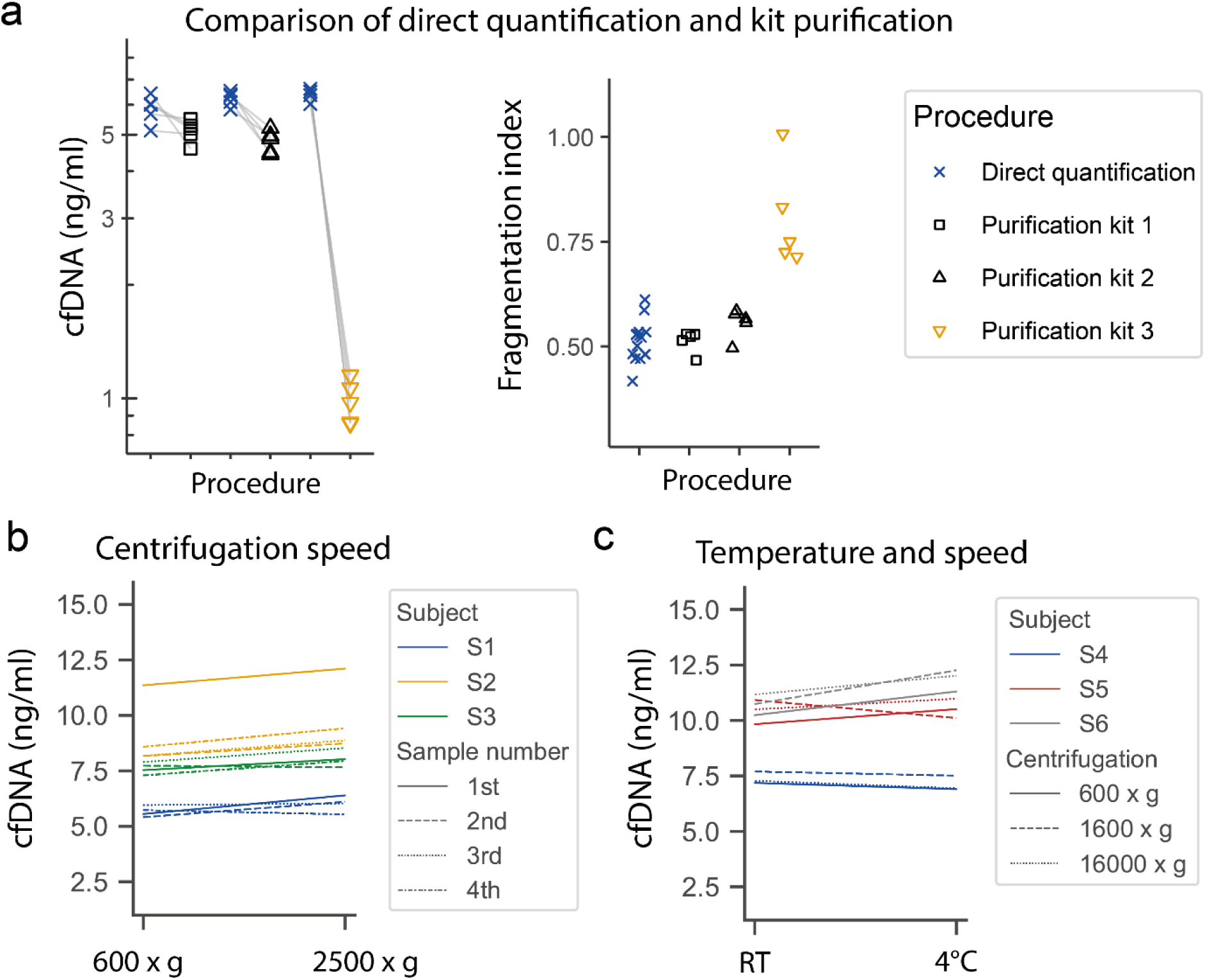
Direct quantification of cfDNA compared to DNA purification and influence of centrifugation speed and temperature on cfDNA concentration. (**a**) Plasma aliquots of the same sample were directly quantified or analyzed after DNA isolation with custom DNA isolation kits, to determine the cfDNA concentration (left) and the fragmentation index (right). (**b**) From different subjects (S1-S3) four samples were consecutively collected in different blood containers, centrifuged at 600 x g or 2500 x g for 10 min and measured with the L1PA2_90bp assay. (**c**) From each subject a single blood sample was collected and centrifuged consecutively at indicated speed at 4°C or room temperature (RT) and measured in duplicate, mean values are illustrated.

### Influence of centrifugation speed or temperature

As shown in Fig. 3b and 3c, centrifugation speed and temperature does not influence the cfDNA concentration in venous plasma. The concentration differences are within the assay precision of the L1PA2_90bp assay. Notably, as shown in Fig. 3b in one subject the first of 4 taken samples shows higher cfDNA levels (independent from centrifugation speed), indicating that the first sample should be discarded.

### Incurred sample reanalysis

During the course of the study ∼17% of the SLE study samples were reanalyzed. The cfDNA concentrations correlate well between first and repeated analysis (r= 0.925, p<0.0001) (Fig. 2b). Among the reanalyzed samples, 5 samples (9.16%) measured with the L1PA2_90bp assay show a difference > ±30% between the initial and the repeated measurement (Fig. 2c). 25.53% of the reanalyzed samples show a difference > ±30% between first and repeated measurement in the L1PA2_222bp assay.

### cfDNA measurement in study samples

A total of 280 study samples were measured with each of the assays. Seven (2.5%) of the samples measured with the L1PA2_90bp, and 21 (7.5%) measured with the L1PA2_222bp assay showed repeatedly high SD of Cq values and were declared to be not measurable.

### Kinetics of cfDNA after exercise

The estimated means of the SLE patients’ venous cfDNA levels increased significantly ∼ 2.1 fold from 13.9 ng/ml (95% CI: 10.4–18.8) to 29.6 ng/ml (95% CI: 22.0–39.9) and decreased to 14.1 ng/ml (95% CI: 10.5–19.9) 90 min after walking exercise (Fig. 4c). Similarly, the capillary samples increased significantly ∼ 2.2 fold from 11.1 ng/ml (95% CI: 8.0–15.4) to 24.8 ng/ml (95% CI: 17.9–34.4) and decreased to 13.6 ng/ml (95%CI: 10.0–18.4). After 90 min the concentrations declined to PRE exercise values again. A detailed results table for the multiple comparisons is given in Supplementary Table S8 and S9. Fig. 4a and 4b display the differences of cfDNA levels from the PRE value.

**Figure 4.**
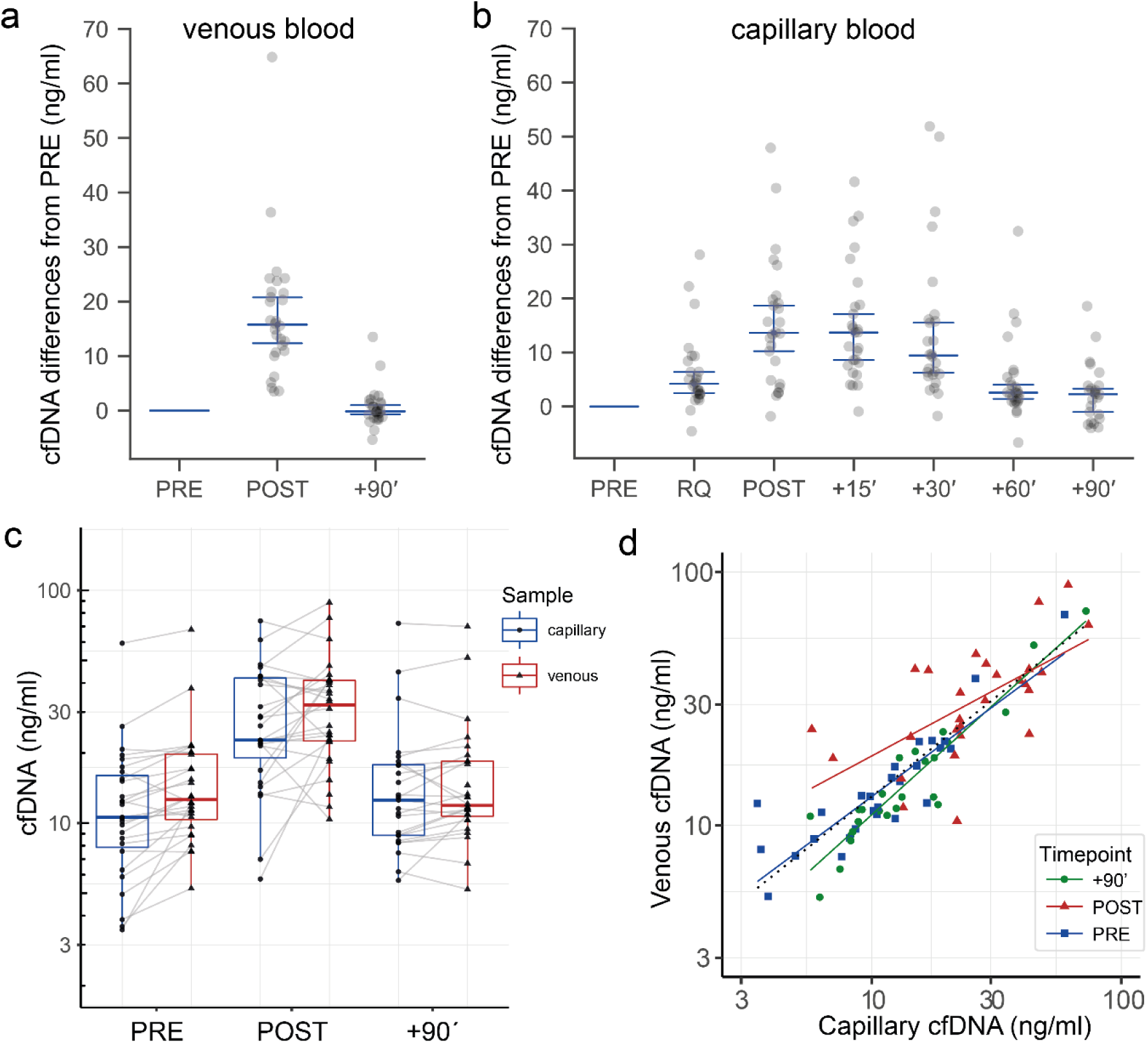
Kinetics of venous and capillary cfDNA measured with the L1PA2_90bp assay before and after exercise. Differences of cfDNA from PRE samples during and after exercise in venous plasma samples (**a**) and capillary plasma samples (**b**) (median and 95% CI are illustrated). (**c**) Boxplots of cfDNA concentration in capillary and venous plasma samples. (**d**) Correlation between venous and capillary cfDNA concentrations.

The comparison between venous and capillary cfDNA concentrations (Fig. 4d) shows an overall correlation of *R*^*2*^= 0.774, p < 0.0001 (dotted black line). The correlation between venous and capillary samples is higher for the PRE exercise (*R*^*2*^ = 0.933, p < 0.0001), and +90 min samples (*R*^*2*^ = 0.974, p < 0.0001) compared to POST exercise samples (ρ = 0.726, p = 0.0002).

As indicated in Fig. 5a cfDNA remains increased until 30 min post exercise. The linear regression across all time points (black line) indicates a half-life of ∼30 min (slope = -0.167, intercept = 16.87). Between 30 min post exercise and 60 min post exercise the cfDNA concentrations show the highest decay rate. The linear regression between +30 and +60 min indicates a half-life of about ∼15 min (red regression line, slope = -0.315, intercept = 23.61). The fragmentation index, calculated as the quotient of the concentrations of the longer L1PA2_222bp fragments and the shorter L1PA2_90bp fragments, increases from 0.33 ± 0.23 ng/ml at the PRE time point to 0.35 ± 0.23 ng/ml post exercise, and decreases to 0.32 ± 0.26 ng/ml after 90 min (Fig. 5b).

**Figure 5.**
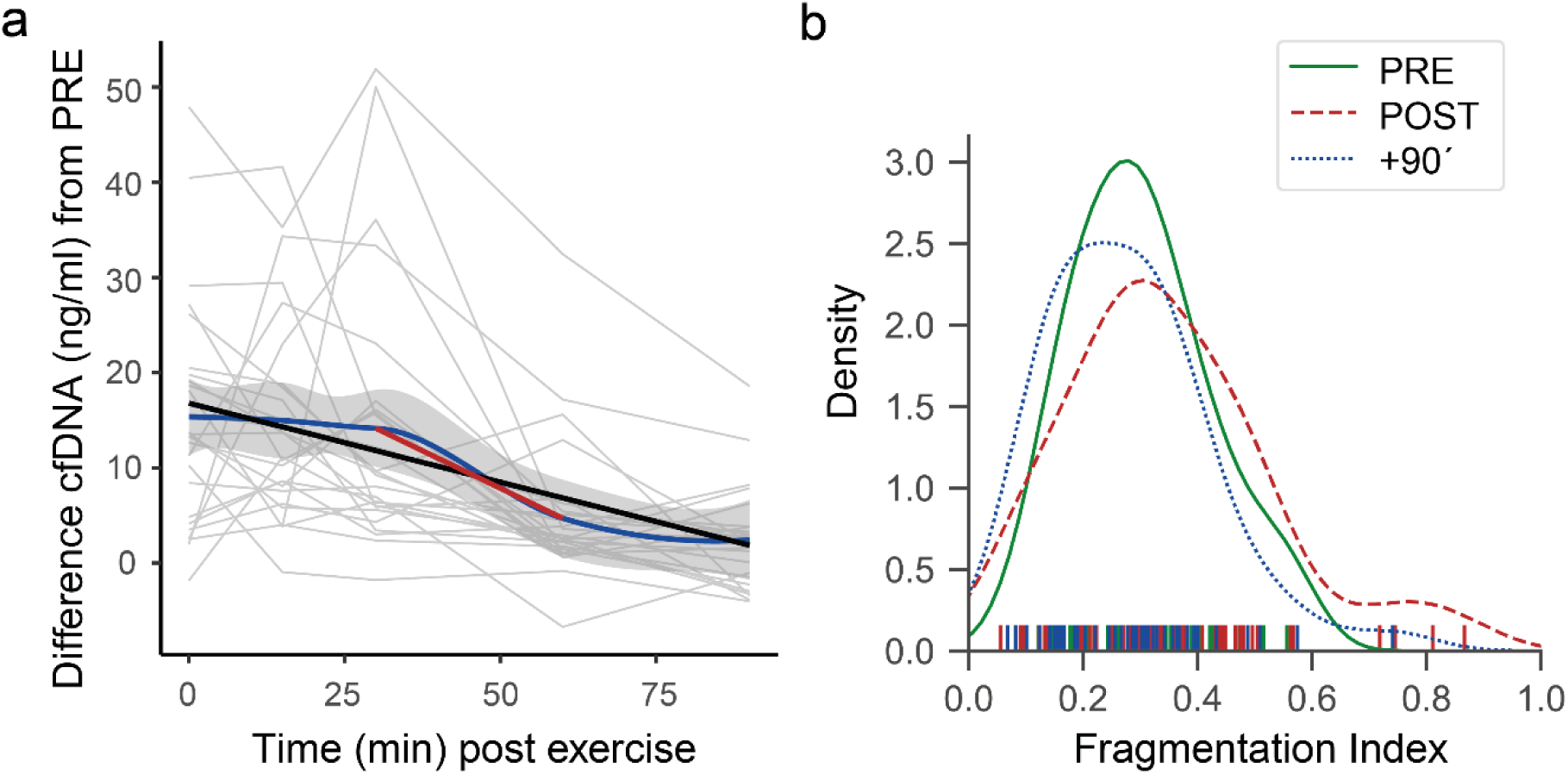
cfDNA decay and fragmentation index. (**a**) The ‘loess’ regression line (blue, with 95% CI in grey) indicates that levels of cfDNA remain increased until 30 min post exercise and decrease most strongly between +30’ and +60’ (red linear regression line). (**b**) Distribution of the fragmentation index of all study samples.

## Discussion

During the last years, cfDNA has been increasingly studied for its prognostic or diagnostic potential in various clinical fields^2,32,33^. A prerequisite to use cfDNA levels for monitoring applications is to ensure reliable and reproducible quantification^34^. The ISO guideline ISO 20395:2019 specifies the requirements for evaluating the performance of quantification methods for nucleic acid target sequences for qPCR and dPCR experiments^18^. The guideline provides a profound background to validate qPCR assays, comprising the MIQE and relevant CLSI guidelines. To compare the cfDNA concentrations of the SLE study samples over multiple runs, we implemented an inter-plate calibration procedure which allows reliable comparison of cfDNA concentrations between different qPCR runs. To ensure the reproducibility of the direct quantification method in plasma of SLE patient who receive medication we applied incurred sample re-analysis, to verify the concentration of cfDNA^35^.

The, established L1PA2_90bp and L1PA2_222bp qPCR assays enable reliable direct quantification of cfDNA in minute amounts of plasma samples. A unique advantage of the assays is that the plasma samples do not need to be purified. A step which is time and cost consuming and can introduce errors in the quantification process. As shown in Fig. 3a, especially DNA isolation kits, which are not specifically established to isolate cfDNA from plasma can lead to reduced cfDNA concentrations and the loss of short DNA fragments^15,36^. Importantly, our assay could be used as a standard to control for DNA losses and to estimate the integrity of the isolated DNA samples.

At the time, there is no defined number of replicates to determine LOQ, whereas a minimum of 10 replicates in total per concentration is recommended^18^. For the precision studies, pooled PRE and POST exercise plasma were used representing low and high copy number samples. The CV for all samples, were below 10%, indicating sufficient precision to clearly discriminate the effects of exercise. To determine the relative repeatability and relative run-to-run variation standard deviations, duplicates of the low and high copy plasma pools were measured in 10 consecutive qPCR runs. All precision estimations were below 12.1%.

Whenever samples are not analyzed in the same run, it is important to take inter-run calibration into account. For qPCR assays this is of unique importance because the relationship between quantification cycle and relative quantity is run dependent^37^. The baseline correction of each qPCR run is depending on the total number of samples and the fluorescence intensity^37^. Here, we provided experimental proof that the use of two inter-run calibrators reduced the intermediate precision between 8.07% and 16.29%.

A number of pre-analytical variables can affect the results of cfDNA measurement^15,38^. As reviewed by Ungerer et al.^15^ the influence of centrifugation speed is still discussed controversially, whereas a second centrifugation step is recommended^38^. Our results indicate that centrifugation at 600 x g compared to 2500 or 16.000 x g does not relevantly influence cfDNA concentrations. The detectable differences were within the assay precision. Notably, we avoided untarred centrifuges, and disturbances of the cell pellet during pipetting. Similar to other studies we did not detect cfDNA changes after prolonged storage of EDTA whole blood samples to several hours^39,40^.

According to the simpler, and less invasive sampling technique, capillary plasma samples are a reasonable sample source, which can be collected more frequently. The comparison of venous and capillary samples shows high congruence. Notably, repeatedly collected capillary samples show a higher variance of cfDNA concentrations compared to venous samples, which is not related to assay imprecision (as indicated in Supplementary Fig. S7). The reason is not clarified in detail, and the intra-individual variance needs further systematic evaluation.

Studies indicate that SLE patients show higher cfDNA concentrations compared to the healthy population, and that cfDNA concentrations are related to disease activity^12,41^. Exercise is recommended for the treatment of SLE patients^42^. However, the kinetics of cfDNA after exercise has not been studied. After walking until exhaustion, the studied SLE patients showed lower fold-changes compared to healthy subjects, which is likely related to exercise intensity, time until exhaustion, and total energy expenditure^7,14,43^. cfDNA concentrations normalized 60-90 min after exercise in most of the patients (Fig. 4 and Fig. 5), showing no significant differences from the PRE cfDNA values (Supplementary Tables S8 and S8). Since elevated cfDNA concentrations have been discussed to possibly trigger enhanced inflammation^44^, or the production of anti-ds-DNA antibodies^5^, low increases and a rapid decrease are positive aspects of cfDNA kinetics in SLE patients. With the established time and costs efficient assay we determined the cfDNA kinetics in exercising SLE patients for the first time, underlining that exercise does not has a negative impact on the levels of cfDNA.

## Data Availability

All data are available from the corresponding author on reasonable request.

## Data availability

All data are available from the corresponding author on reasonable request.

