## Supplemental Material for "Validating quantitative PCR assays for cell-free DNA detection without DNA extraction: Exercise induced kinetics in systemic lupus erythematosus patients"

**Supplementary Table S1:** Utilized blood collection devices, plastic ware, pipettes and chemicals for the qPCR assays.

| **Blood collection devices, Plastic ware, and pipetts** | **Vendor** | **Cat #** |
|---|---|---|
| Safety-Lancet: Safety-Lanzette Extra 21G | SARSTEDT | 85.1016 |
| Microvette® CB 300 K2E | SARSTEDT | 16.444 |
| S-Monovette® 7.5 ml, K3 EDTA, 92x15 mm | SARSTEDT | 1605001 |
| Safety-Multifly®-Needle 21G tube 80mm | SARSTEDT | 85.1638.203 |
| Filter tips "Avantgμard" 10 µl XL, sterile, surface optimized, with extremely fine tip | Axon Labortechnik | 23213 |
| FrameStar® 384-well PCR plates (white wells) | Bio-Budget | 34-480LC-0384 |
| AMPLIseal™ Transparent adhesive sealer for RT PCR | Greiner Bio-One | 676040 |
| Reaction tubes 0.2 | Greiner Bio-One | 683201 |
| Reaction tubes 0.5 | Greiner Bio-One | 667201 |
| Reaction tubes 1.5 ml | Greiner Bio-One | 616201 |
| ErgoOne® Single-Channel Pipette, 0.1 – 2.5 µl | STARLAB | S7100-0125 |
| Eppendorf Reference® 2 Ein-Kanal, fix, 20 µL | eppendorf | EP4921000060 |

| **Chemicals for qPCR** | **Vendor** | **Cat #** |
|---|---|---|
| UltraPure™ DNase/RNase-Free Distilled Water | Invitrogen™ | 10977049 |
| VELOCITY DNA Polymerase (500 Units) | BioCat | BIO-21099-BL |
| 5x Hi-Fi Buffer (provided with polymerase) | BioCat | |
| dNTP Mix (40mM Final Conc.) | BioCat | BIO-39043-BL |
| SYBR Green nucleic acid gel stain 10,000 x | Sigma | C90M1158 |

**Supplementary Table S2:** Sequence of the custom made L1PA2 DNA fragment (GRCh38/hg38_ chr4:68,085,016-68,085,410 / size = 395 bp / strand = +)

| Sequence (5´ → 3´) |
|---|
| gaattcAGAATGATGATTTCCAATTTCATCCATGTCCCTACAAAGGACATGAACTCATCAT |
| TTTTTATGGCTGCATAGTATTCCATGGTGTATATGTGCCACATTTTCTTAATCCAGTCT |
| ATCATTGTTGGACATTTGGATTGGTTCCAAGTCTTTGCTATTGTGAATAA***TGCCGCAAT*** |
| ***AAACATACGTG***TGCATGTGTCTTTATAGCAGCATGATTTATAGTCATTTGGGTATATAC |
| CCA***GTAATGGGATGGCTGGGTC***AAATGGTATCTCTAGTTCTAGATCCCTGAGGAATC |
| GCCACACTGACTTCCACAATGGTTGAACTAGTTTACAGTCCCACCAACAGTGTAAAAG |
| TGTTCCTATTTCTCCACAT***CCTCTCCAGCACCTGTTGTT***TCCTGACTTgaattc |

The binding site for the L1PA2_fw primer is highlighted in grey. The binding sites for the reverse primers of the L1PA2_90bp and L1PA2_222bp assays are highlighted in blue and green, respectively.

**Supplementary Table S3:** Primer sequences

| L1PA2 PCR Primers [0.14 µmol/PCR] | Sequence in 5' → 3' | Amplicon size | Annealing temperature |
|---|---|---|---|
| L1PA2_fw | TGCCGCAATAAACATACGTG | | 60.4 °C |
| L1PA2_90bp_rv | GACCCAGCCATCCCATTAC | 90 bp | 61.1 °C |
| L1PA2_222bp_rv | AACAACAGGTGCTGGAGAGG | 222 bp | 63.4 °C |

**Supplementary Figure S4:** Distribution of the hits per chromosome in the human genome (GRCh38/hg38)

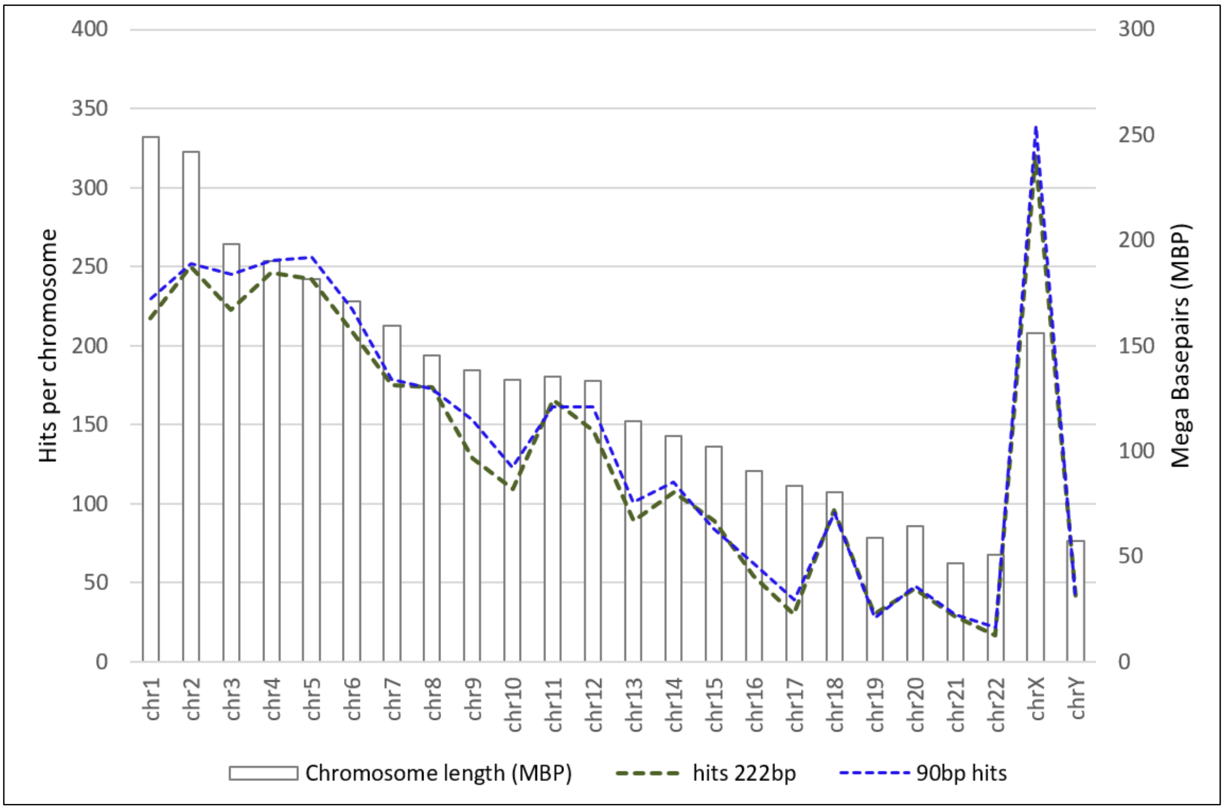

**Supplementary Table S5:**

Predicted targets from the *UCSC In-Silico PCR* (length and number) for the L1PA2_fw and L1PA2_222bp_rv primer pair in GRCh38/hg38.

| **L1PA2_90bp** | |
|---|---|
| Sequence length | Hits in the Genome |
| 90bp | 3338 |
| 89bp | 28 |
| 91bp | 14 |
| 92bp | 7 |
| 88bp | 5 |
| 87bp | 4 |
| 68bp | 2 |
| 79bp | 2 |
| 321bp | 2 |
| 96bp | 2 |
| 330bp | 1 |
| 422bp | 1 |
| 94bp | 1 |
| 278bp | 1 |
| 186bp | 1 |
| 76bp | 1 |
| 97bp | 1 |
| 260bp | 1 |
| 86bp | 1 |
| 81bp | 1 |
| 95bp | 1 |
| **Total** | **3416** |

| **L1PA2_222bp** | |
|---|---|
| Sequence length | Hits in the Genome |
| 222bp | 2985 |
| 221bp | 117 |
| 223bp | 35 |
| 220bp | 20 |
| 219bp | 10 |
| 224bp | 10 |
| 226bp | 5 |
| 228bp | 4 |
| 212bp | 4 |
| 227bp | 4 |
| 218bp | 3 |
| 213bp | 2 |
| 210bp | 2 |
| 200bp | 2 |
| 217bp | 2 |
| 209bp | 2 |
| 204bp | 2 |
| 240bp | 2 |
| 230bp | 2 |
| 216bp | 2 |
| 232bp | 2 |
| 197bp | 1 |
| 235bp | 1 |
| 236bp | 1 |
| 225bp | 1 |
| 410bp | 1 |
| 229bp | 1 |
| 215bp | 1 |
| 408bp | 1 |
| 205bp | 1 |
| 392bp | 1 |
| 233bp | 1 |
| 214bp | 1 |
| 242bp | 1 |
| 238bp | 1 |
| 190bp | 1 |
| 211bp | 1 |
| 558bp | 1 |
| 208bp | 1 |
| 203bp | 1 |
| 256bp | 1 |
| **Total** | **3237** |

**Supplementary Fig. S6:** Typical amplification curves of the reference samples and $H_2O$ or mouse plasma NTCs of the L1PA2_90bp assay (A), and corresponding melt curves (B). Typical L1PA2_222bp amplification curves of the reference samples with NTC (C), and corresponding melt curves (D). RFU = relative fluorescence units.

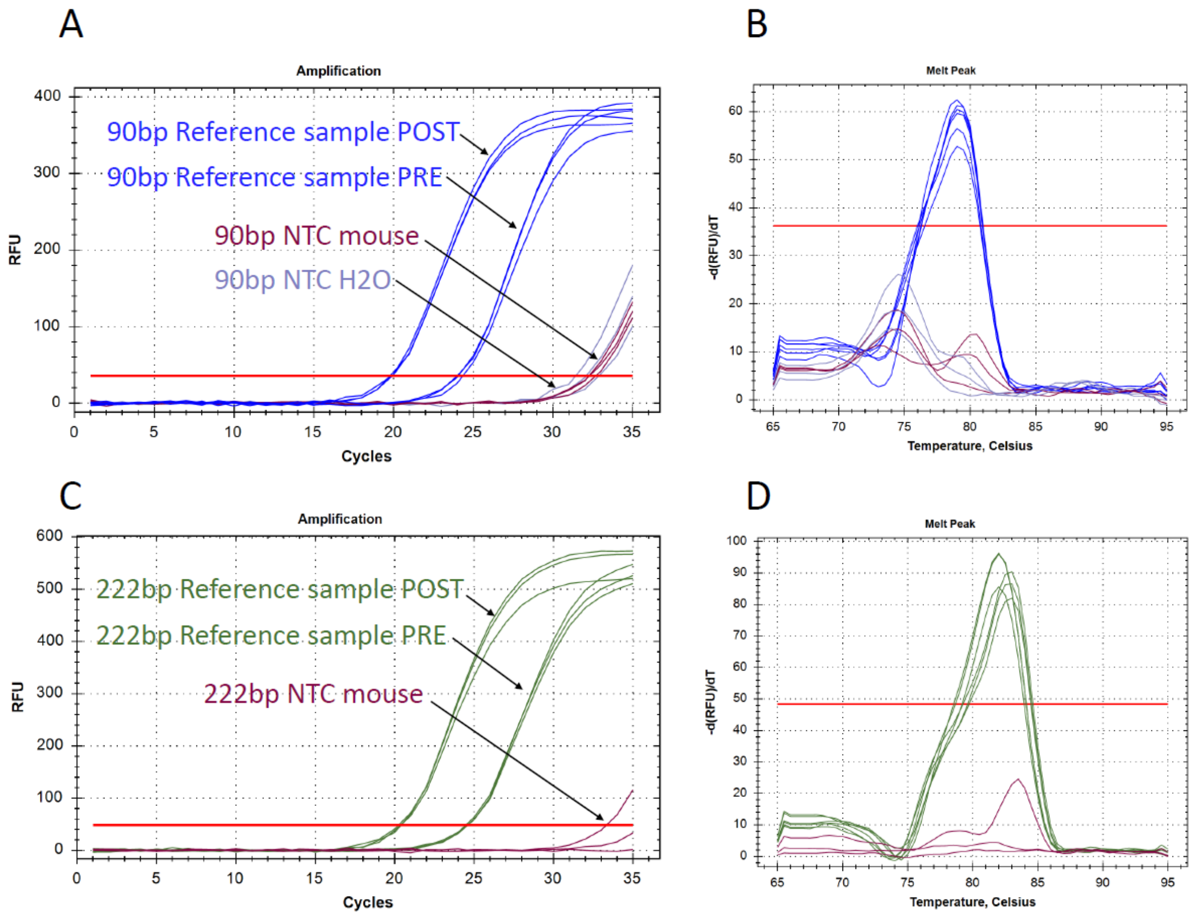


**Supplementary Fig. S7:** cfDNA concentrations after extended storage of blood samples.

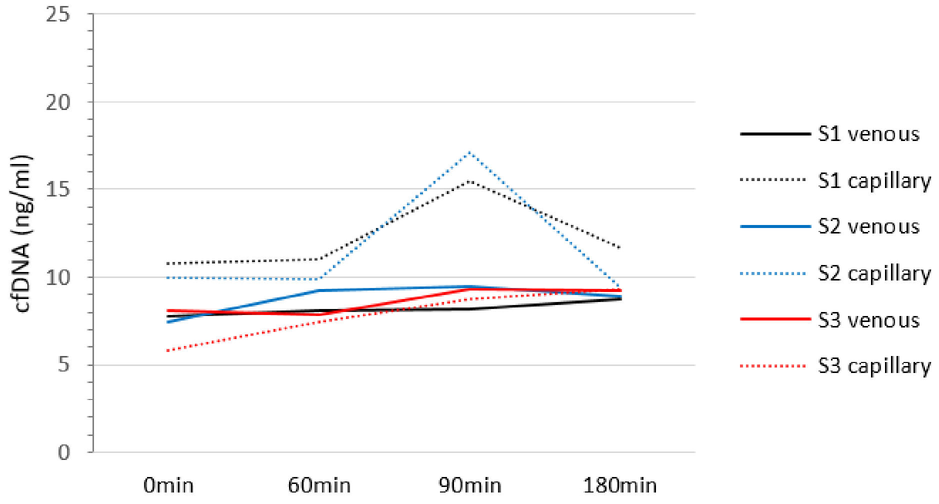


After prolonged storage of the blood samples before centrifugation (60min, 90 min, or 180 min) the cfDNA concentrations do not differ relevantly in three subjects. Capillary cfDNA samples show a higher variance compared to venous samples.

**Supplementary Table S8:** Results of the post-hoc comparisons between capillary and venous samples, related to Linear mixed model fit by REML ['lmerMod'].

Formula: log10(cfDNA_ng_ml) ~ Timepoint * Sample_type + (1 | Subject)

| Timepoint | Sample_type | response | SE | df | lower.CL | upper.CL |
|---|---|---|---|---|---|---|
| +90 min | capillary | 13.6 | 1.49 | 44.9 | 10.0 | 18.4 |
| POST | capillary | 24.8 | 2.69 | 43.0 | 18.3 | 33.4 |
| PRE | capillary | 11.1 | 1.20 | 43.0 | 8.2 | 14.9 |
| +90 min | venous | 14.1 | 1.53 | 42.1 | 10.5 | 19.0 |
| POST | venous | 29.6 | 3.20 | 42.1 | 22.0 | 39.9 |
| PRE | venous | 13.9 | 1.51 | 42.1 | 10.4 | 18.8 |

Degrees-of-freedom method: kenward-roger
Confidence level used: 0.95
Conf-level adjustment: sidak method for 6 estimates
Intervals are back-transformed from the log10 scale

$contrasts

| contrast | estimate | SE | df | t.ratio | p.value |
|---|---|---|---|---|---|
| (+90 min capillary) - POST capillary | -0.26040 | 0.0345 | 130 | -7.544 | <.0001 |
| (+90 min capillary) - PRE capillary | 0.08916 | 0.0345 | 130 | 2.583 | 0.1089 |
| (+90 min capillary) - (+90 min venous) | -0.01662 | 0.0342 | 130 | -0.487 | 0.9966 |
| (+90 min capillary) - POST venous | -0.33834 | 0.0342 | 130 | -9.906 | <.0001 |
| (+90 min capillary) - PRE venous | -0.01128 | 0.0342 | 130 | -0.330 | 0.9995 |
| POST capillary - PRE capillary | 0.34956 | 0.0337 | 130 | 10.367 | <.0001 |
| POST capillary - (+90 min venous) | 0.24378 | 0.0334 | 130 | 7.308 | <.0001 |
| POST capillary - POST venous | -0.07794 | 0.0334 | 130 | -2.337 | 0.1871 |
| POST capillary - PRE venous | 0.24912 | 0.0334 | 130 | 7.468 | <.0001 |
| PRE capillary - (+90 min venous) | -0.10578 | 0.0334 | 130 | -3.171 | 0.0228 |
| PRE capillary - POST venous | -0.42750 | 0.0334 | 130 | -12.816 | <.0001 |
| PRE capillary - PRE venous | -0.10044 | 0.0334 | 130 | -3.011 | 0.0362 |
| (+90 min venous) - POST venous | -0.32172 | 0.0330 | 130 | -9.750 | <.0001 |
| (+90 min venous) - PRE venous | 0.00534 | 0.0330 | 130 | 0.162 | 1.0000 |
| POST venous - PRE venous | 0.32706 | 0.0330 | 130 | 9.912 | <.0001 |

Note: contrasts are still on the log10 scale
Degrees-of-freedom method: kenward-roger
P value adjustment: tukey method for comparing a family of 6 estimates

**Supplementary Table S9:** Results of the post-hoc comparisons between capillary samples at all timepoints, related to Linear mixed model fit by REML ['lmerMod'].

Formula: log10(cfDNA_ng_ml) ~ Timepoint + (1 | Subject)

| Timepoint | response | SE | df | lower.CL | upper.CL |
|---|---|---|---|---|---|
| +15 min | 24.7 | 2.87 | 45.8 | 17.83 | 34.2 |
| +30 min | 22.5 | 2.63 | 46.9 | 16.18 | 31.2 |
| +60 min | 15.1 | 1.76 | 46.8 | 10.86 | 20.9 |
| +90 min | 13.6 | 1.61 | 49.1 | 9.76 | 18.9 |
| POST | 24.8 | 2.90 | 46.8 | 17.87 | 34.4 |
| PRE | 11.1 | 1.30 | 46.8 | 8.00 | 15.4 |
| RQ | 16.5 | 1.92 | 45.8 | 11.92 | 22.9 |

Degrees-of-freedom method: kenward-roger
Confidence level used: 0.95
Conf-level adjustment: sidak method for 7 estimates
Intervals are back-transformed from the log10 scale

$contrasts

| contrast | estimate | SE | df | t.ratio | p.value |
|---|---|---|---|---|---|
| (+15 min) - (+30 min) | 0.04137 | 0.0382 | 155 | 1.082 | 0.9325 |
| (+15 min) - (+60 min) | 0.21461 | 0.0382 | 155 | 5.616 | <.0001 |
| (+15 min) - (+90 min) | 0.25918 | 0.0391 | 155 | 6.627 | <.0001 |
| (+15 min) - POST | -0.00167 | 0.0382 | 155 | -0.044 | 1.0000 |
| (+15 min) - PRE | 0.34729 | 0.0382 | 155 | 9.088 | <.0001 |
| (+15 min) - RQ | 0.17489 | 0.0378 | 155 | 4.626 | 0.0002 |
| (+30 min) - (+60 min) | 0.17325 | 0.0386 | 155 | 4.485 | 0.0003 |
| (+30 min) - (+90 min) | 0.21781 | 0.0394 | 155 | 5.530 | <.0001 |
| (+30 min) - POST | -0.04303 | 0.0386 | 155 | -1.114 | 0.9230 |
| (+30 min) - PRE | 0.30593 | 0.0386 | 155 | 7.920 | <.0001 |
| (+30 min) - RQ | 0.13352 | 0.0382 | 155 | 3.493 | 0.0109 |
| (+60 min) - (+90 min) | 0.04456 | 0.0395 | 155 | 1.128 | 0.9187 |
| (+60 min) - POST | -0.21628 | 0.0386 | 155 | -5.601 | <.0001 |
| (+60 min) - PRE | 0.13268 | 0.0386 | 155 | 3.436 | 0.0131 |
| (+60 min) - RQ | -0.03972 | 0.0382 | 155 | -1.040 | 0.9440 |
| (+90 min) - POST | -0.26084 | 0.0395 | 155 | -6.601 | <.0001 |
| (+90 min) - PRE | 0.08812 | 0.0395 | 155 | 2.230 | 0.2858 |
| (+90 min) - RQ | -0.08429 | 0.0391 | 155 | -2.155 | 0.3263 |
| POST - PRE | 0.34896 | 0.0386 | 155 | 9.036 | <.0001 |
| POST - RQ | 0.17655 | 0.0382 | 155 | 4.620 | 0.0002 |
| PRE - RQ | -0.17240 | 0.0382 | 155 | -4.512 | 0.0003 |

Note: contrasts are still on the log10 scale
Degrees-of-freedom method: kenward-roger
P value adjustment: tukey method for comparing a family of 7 estimates
